# Clinical management and mortality among COVID-19 cases in sub-Saharan Africa: A retrospective study from Burkina Faso and simulated case analysis

**DOI:** 10.1101/2020.06.04.20119784

**Authors:** Laura Skrip, Karim Derra, Mikaila Kaboré, Navideh Noori, Adama Gansané, Innocent Valéa, Halidou Tinto, Bicaba W. Brice, Mollie Van Gordon, Brittany Hagedorn, Hervé Hien, Benjamin M. Althouse, Edward A. Wenger, André Lin Ouédraogo

## Abstract

**Background:** Absolute numbers of COVID-19 cases and deaths reported to date in the sub-Saharan Africa (SSA) region have been significantly lower than those across the Americas, Asia, and Europe. As a result, there has been limited information about the demographic and clinical characteristics of deceased cases in the region, as well as the impacts of different case management strategies.

**Methods:** Data from deceased cases reported across SSA through May 10, 2020 and from hospitalized cases in Burkina Faso through April 15, 2020 were analyzed. Demographic, epidemiological, and clinical information on deceased cases in SSA was derived through a line-list of publicly available information and, for cases in Burkina Faso, from aggregate records at the Center Hospitalier Universitaire de Tengandogo in Ouagadougou. A synthetic case population was derived probabilistically using distributions of age, sex, and underlying conditions from populations of West African countries to assess individual risk factors and treatment effect sizes. Logistic regression analysis was conducted to evaluate the adjusted odds of survival for patients receiving oxygen therapy or convalescent plasma, based on therapeutic effectiveness observed for other respiratory illnesses.

**Results:** Across SSA, deceased cases for which demographic data are available have been predominantly male (63/103, 61.2%) and over 50 years of age (59/75, 78.7%). In Burkina Faso, specifically, the majority of deceased cases either did not seek care at all or were hospitalized for a single day (59.4%, 19/32); hypertension and diabetes were often reported as underlying conditions. After adjustment for sex, age, and underlying conditions in the synthetic case population, the odds of mortality for cases not receiving oxygen therapy was significantly higher than those receiving oxygen, such as due to disruptions to standard care (OR: 2.07; 95% CI: 1.56 – 2.75). Cases receiving convalescent plasma had 50% reduced odds of mortality than those who did not (95% CI: 0.24 – 0.93).

**Conclusion:** Investment in sustainable production and maintenance of supplies for oxygen therapy, along with messaging around early and appropriate use for healthcare providers, caregivers, and patients could reduce COVID-19 deaths in SSA. Further investigation into convalescent plasma is warranted, as data on its effectiveness specifically in treating COVID-19 becomes available. The success of supportive or curative clinical interventions will depend on earlier treatment seeking, such that community engagement and risk communication will be critical components of the response.

## Background

At two months post-introduction into the region, countries across sub-Saharan Africa (SSA) have been reporting significantly fewer cases of SARS-CoV-2 (COVID-19) than higher income countries and some low and middle income countries (LMICs) on other continents.^1^ As a result, the majority of evidence to date reflects disease progression, hospitalization rates, and deaths among cases in more developed and higher transmission settings across Europe, North America,^2^ and Asia.^3,4^ While absolute case and death counts are lower in SSA, individual countries are reporting high case fatality ratios (CFRs) and available information warrants investigation into mortality reduction strategies. It has been hypothesized that the SSA region is vulnerable to high rates of COVID-19 deaths given limited healthcare capacity to manage critical cases and poor detection leading to delayed care-seeking.^5-7^

Across SSA, healthcare-related resource constraints range from inadequate supplies of medical equipment to low per capita capacity for isolation and treatment. A recent report estimated that the entire African continent has 1% of the ventilator capacity of the United States.^8^ Furthermore, countries have reported between one and 36 hospital beds per 10,000 capita.^9^ These constraints have been at odds with policies to quarantine presumptive cases in precautionary isolation centers^10,11^ or isolate all positive cases in dedicated hospitals as a control strategy.^12^ Moreover, messaging around the lack of effective treatment for COVID-19 or unavailability of resources may be deterring early care-seeking among individuals who develop symptoms and could benefit from supportive care. Given systemic vulnerabilities in health system capacity, it is critical to investigate and invest in lower cost and readily scalable interventions, including some already with precedent for reducing the morbidity and mortality associated with respiratory and/or viral infections. Notably, oxygen therapy^13^ and use of blood-related therapies^14^ hold potential for not only managing COVID-19 cases but also treating other conditions prevalent or emerging in LMICs.

Burkina Faso was among the first countries in SSA to report COVID-19 cases. The first case of COVID-19 in Burkina Faso was confirmed on March 9, 2020. Since then, several containment measures have been adopted by the government of Burkina Faso, including social distancing, closure of airports and land borders, quarantine of the affected cities and the mandatory use of masks (Figure 1). Such efforts may be suppressing transmission, yet the country is reporting among the highest numbers of deaths regionally.

**Figure 1.**
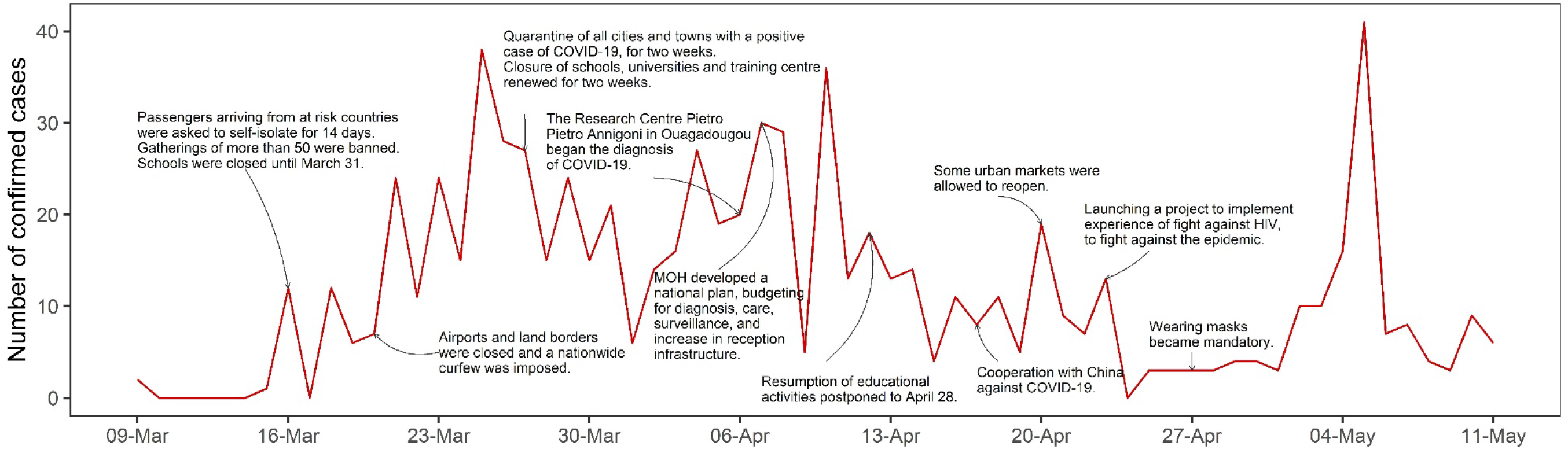
COVID-19 epidemic curve (weekly cases in red) and timeline of control efforts in Burkina Faso, 3/9/2020-5/11/2020. The first case in Burkina Faso was detected on March 9, 2020. The Centre for Research Centre Pietro Annigoni (CERBA) in Ouagadougou, initiated diagnosis of COVID-19 on April 6th, and as of April 21, the Centre has analyzed 460 samples. Two more laboratories in Ouagadougou, The Yalgado Hospital Centre and the National Public Health Laboratory, also analyze the same samples to ensure the quality of the results. On April 7th, the Minister of Health in Burkina Faso presented a national plan to respond to COVID-19 pandemic. In this plan, budgets are specifically allocated to diagnosis, surveillance, infection prevention and control, patient safety, research, and developing remote quick intervention equipment.

COVID-19 cases in Africa currently comprise approximately 1% of the reported global burden.^15^ Low levels of detection, partially due to insufficient testing capacity and laboratory supplies, and the lack of published data from individual African countries challenge understanding of the true scale of infection as well as study of potential risk factors and intervention approaches.^7^ Here we seek to leverage existing, yet minimal, data and generate insight into the impact of potentially lower cost and more logistically feasible approaches to case management in the LMIC context, where care-seeking may be affected by the perceived lack of treatment options. We provide an overview of demographic and clinical characteristics of deceased cases across SSA overall and in Burkina Faso specifically. Trends in mode of detection and care-seeking among all cases were compared to trends among cases who ultimately died of the disease. Mitigation strategies to prevent current excess mortality while also preparing for future treatment opportunities such as the use of blood-related products, were investigated through statistical analysis of a synthetic case population matched to demographic and clinical characteristics of cases in West Africa.

## Methods

### Data

Data from publicly available sources were line-listed to assess epidemiological and clinical characteristics of deceased COVID-19 cases across the entire SSA region. Specifically, data on sex, age, underlying conditions, and mode of detection were identified for deceased cases as previously done for a line list being curated for all cases in SSA.^7^ Additionally, data on age, sex, and time in hospital were recorded for 49 deceased cases reported as of May 10, 2020 and 163 hospitalized cases reported as of April 15, 2020, from the Center Hospitalier Universitaire (CHU) de Tengandogo in Ouagadougou, Burkina Faso. Data are presented as frequencies and percentages. Age distributions between hospitalized and deceased cases were compared using a chi-squared test.

### Data simulation and statistical analysis

We generated a synthetic case study population of individual COVID-19 cases using multinomial distribution sampling.^16-18^ Specifically, for a fixed stratum n and a vector of proportions π=(π1,π2,…,πk), we sampled X ~ Mult (n, π) to attain a sample representative of characteristics (age, gender, comorbidity, disease outcome) in the population. The base uninfected population was randomly drawn using demographics as reported by census data to account for age and gender strata in Burkina Faso.^19^ Age-adjusted distributions of non-communicable diseases *(i.e*., hypertension and diabetes) in West African populations were used to randomly assign underlying conditions to individuals.^20,21^ In the absence of age data for all confirmed cases in Burkina Faso, COVID-19 infection was randomly assigned to individuals following the age-binned distribution of cases in Senegal^22^ assuming that the infection dynamics in the two countries are similar. Age distributions of COVID-19 hospitalization and mortality rates were based on data from the CHU in Burkina Faso. Hospitalization and mortality rates in younger and elderly ages were assumed to vary 100% and 50%, respectively, to address uncertainty in measurement and propagate it using non-linear combinations estimates of errors.

To evaluate COVID-19 case fatality rates in the absence of oxygen therapy, we used adjusted odds ratios as increased risk of death by hypoxia (hypoxia is defined with a cut-off for oxygen saturation rate SpO2 < 90%) in patients of Acute Lower Respiratory Infections^23^ and brain injury^24^ to simulate a binomial distribution of oxygen treatment and non-treatment as described in the contingency Table (Supplementary Table 1).

To consider COVID-19 case fatality rates reduction in the presence of convalescent plasma therapy, we simulated a convalescent plasma treatment study in our hypothetical inpatient population. Given the current lack of data on anti-COVID-19 convalescent plasma treatment, we assumed an absolute risk difference in mortality of 10.5% between treatment and non-treatment groups considering that convalescent plasma during the present COVID-19 pandemic is two-fold less likely to reduce mortality than convalescent plasma did during the 1918 Spanish flu pandemic (21% [95% CI, 15% to 27%] in Luke et al.^25^). Sample sizes distribution to meet a 10.5% risk difference in mortality between groups is described in Supplementary Table 1 and hospitalized patients were randomly assigned to the groups.

Associations between the odds of survival and patient characteristics (age, gender, comorbidity), immunotreatment, and health care capacity (oxygen-therapy) on COVID-19 mortality were estimated using logistic regression. The adjusted odds ratio for survival among cases receiving intervention (oxygen therapy or convalescent plasma) versus those not receiving the intervention were calculated with 95% confidence intervals.

### Ethics statement

All data presented here were derived from publicly available sources or provided by CHU as part of the national epidemic response program. The data from CHU were reported in aggregate, as frequency distributions across 10-year age bins, by sex, or length-of-stay in hospitals. The study was therefore not subject to review by an ethics board.

## Results

### Data description

As of May 10, 2020, 976 deaths had been reported out of the 39,104 confirmed COVID-19 cases across 48 countries in the SSA region (regional CFR: 2.5%).^15^ Line-listed, country-reported data on age and/or sex were available for 75 and 103 deceased cases, respectively. Deceased cases were majority male (63/103, 61.2%) and a median 62 years old (range: 28 days - 89 years) (Figure 2). Over 78% of deaths were reported among individuals aged 50 years or older (59/75, 78.7%). For those cases with information available, the majority of deaths were among individuals who self-presented to facilities or were tested post-mortem (20/30, 66.7%), rather than among cases who were being followed as part of active monitoring initiatives.

**Figure 2.**
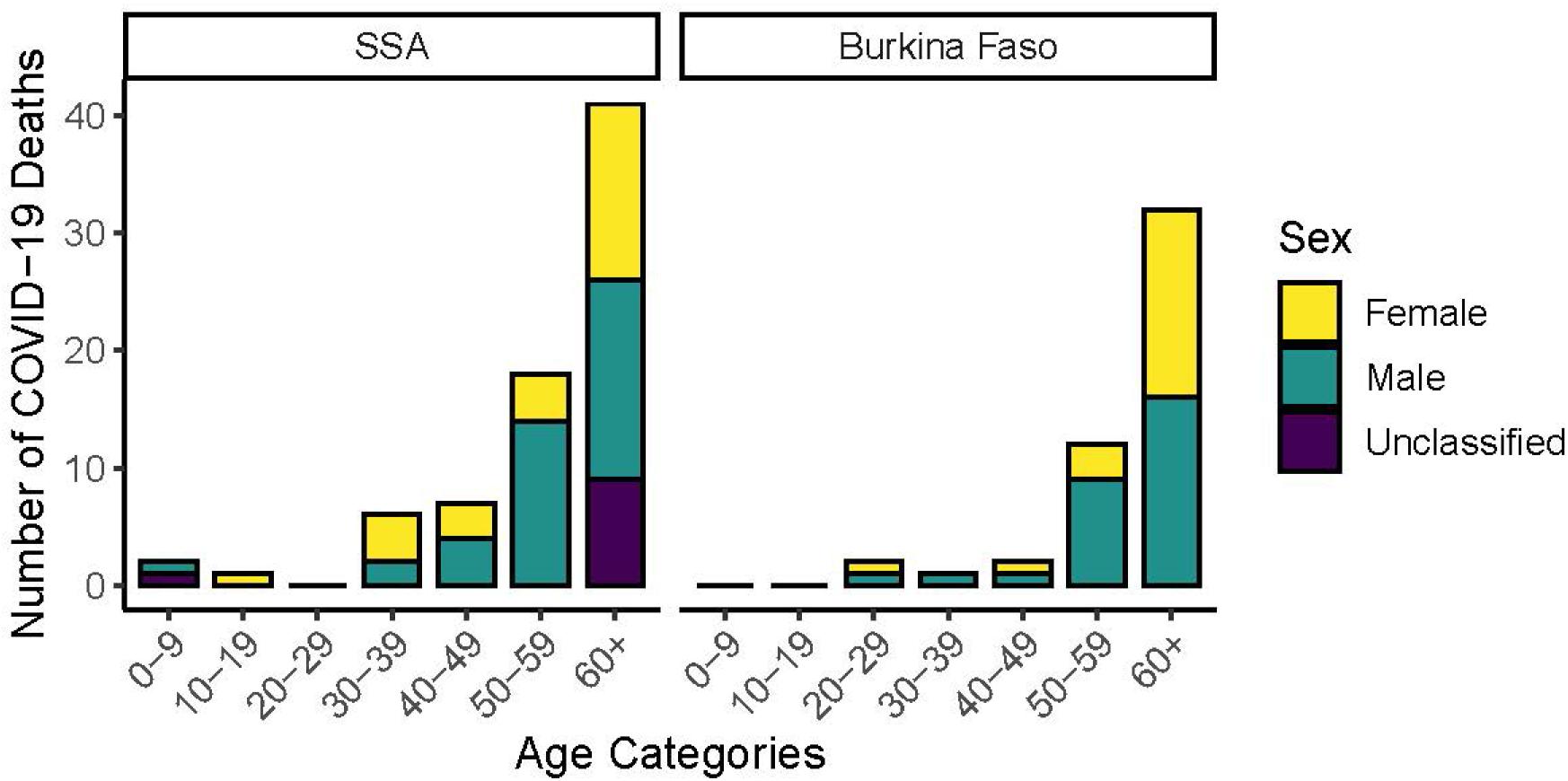
Distribution of sex and age among reported deceased COVID-19 cases in sub-Saharan Africa (SSA) overall (excluding Burkina Faso), and in Burkina Faso.

In Burkina Faso specifically, 49 deaths occurred among 751 detected cases, as of May 10. Data for analysis were available for the 49 deaths through May 10, 2020 and for hospitalized cases (n=163) through April 15, 2020. The majority of deceased cases were male (28/49, 57.1%) and nearly all deaths occurred among individuals aged 50 and above (30/33, 90.9%) (Figure 2). Underlying conditions were prevalent among deceased cases. Nearly half had a history of hypertension (15/33, 45.5%); diabetes (7/33, 21.2%), or other cardiovascular or pulmonary conditions, such as stroke, embolism or cardiopathy (3/33, 9.1%).

Oxygen therapy was most frequently used to treat hospitalized COVID-19 cases before their deaths (27/33, 81.8%) (Figure 3A). Pulsometer values indicate that at time of death, 90% of clinical cases presented with a median oxygen level of 61 (IQR, 45-82) which is below normal value of 90. Clinical treatment also routinely included administration of antimicrobials (Ceftriaxone, Chloroquine, Azithromycin) and fever-reducers (paracetamol). Only two (2/33, 6.1%) critical patients were intubated before their deaths.

**Figure 3.**
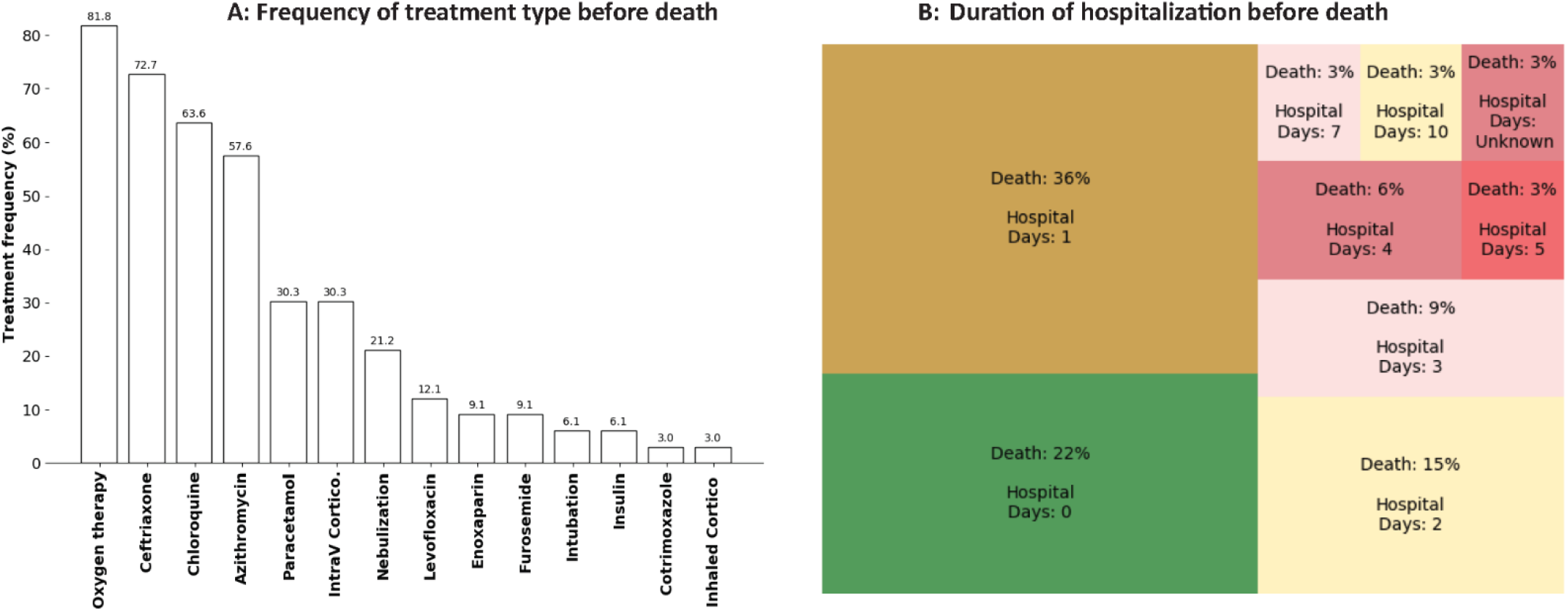
Length of stay in hospital and clinical care administered for 33 COVID-19 cases seeking care at CHU de Tengandogo before death in Burkina Faso

Prior to death, the majority of individuals either did not seek care or were hospitalized for only a short duration: 21.9% (7/32) of cases hospitalized for less than one day or not at all, and 37.5% (12/32) and 15.6% (5/32) hospitalized for one or two days, respectively, before death (Figure 3B). The age distribution of deceased cases differed from that of all hospitalized cases, with the proportion of hospitalized confirmed cases under 50 years of age (56/163, 34.4%) being significantly higher than the proportion of deceased cases under 50 years (5/49, 10.2%) (p=0.002).

### Synthetic population simulation and analysis

Characteristics of the synthetic case population and distribution of COVID-19 cases are shown in Figure 4. In line with the data from Burkina Faso (Figure 4A-C) and other countries in the region, increased rates of underlying conditions, COVID-19 positivity, inpatient treatment-seeking and death were associated with increasing age. Disruption of oxygen therapy was found to approximately double the case fatality rates across age groups. Introduction of convalescent plasma therapy was associated with reduced COVID-19 case fatality rates irrespective of age (Figure 5). In our case population of 831 individuals, the mortality risk difference for those on treatment with convalescent plasma versus those not on treatment was 10.5% [95% CI, −4% to 25%]).

**Figure 4.**
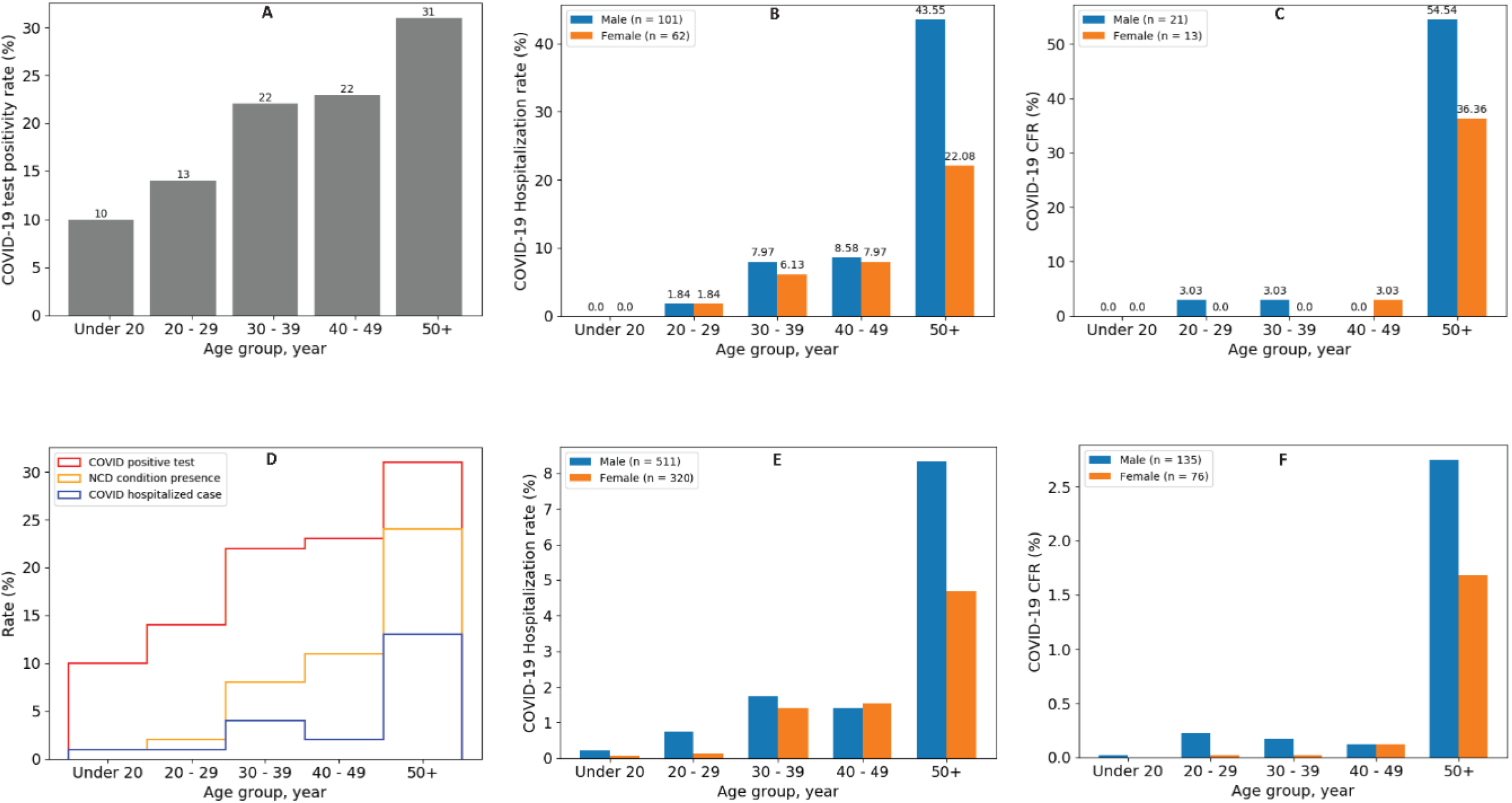
Empirical (Panels A-C) versus simulated (Panels D-F) distributions for age-specific percentages of hospitalizations and underlying conditions in a sample of COVID-19 cases. 10% of the total population was assumed to undergo testing and stratified random sampling was applied to meet test positivity rates as observed in Senegal. Hypothetical population sizes by age groups (under 20, 20-29, 30-39, 40-49 and 50+ years) were estimated for SARS-CoV-2 infections (422, 580, 899, 948, 1257, respectively), hospitalizations (11, 36, 129, 121, 534), and deaths (1, 10, 8, 10, 182).

**Figure 5.**
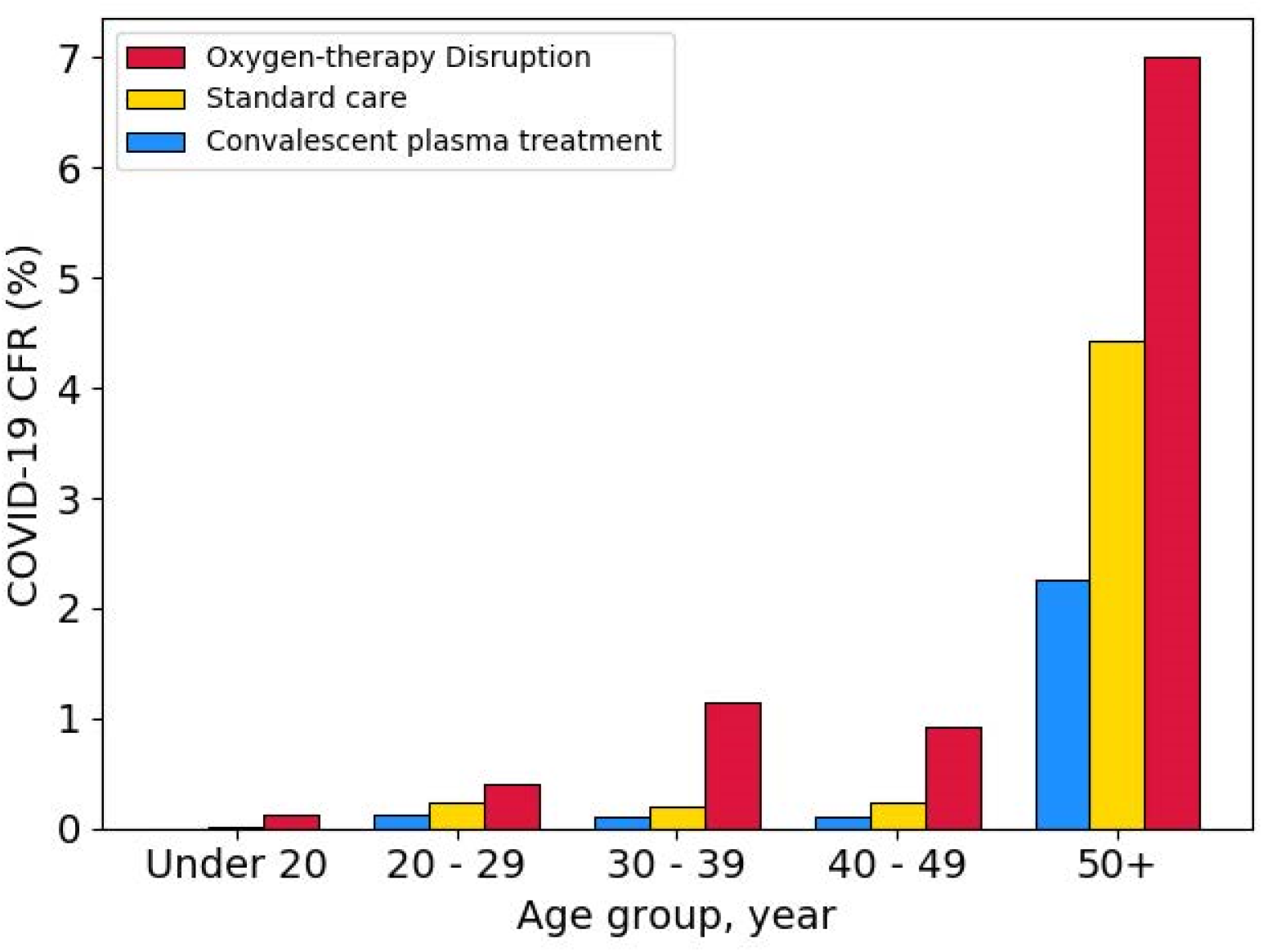
Differential mortality rates modeled as results of convalescent plasma and oxygen therapy effect sizes

Regression results on the impact of individual factors (age, gender, comorbidities), oxygen disruption, and treatment (convalescent plasma) on case fatality rates are shown in Table 1. Age over 50 (AOR = 5.15, 95% CI: 3.34-8.17, p<0.001) was associated with significantly increased odds of mortality in the adjusted analysis. No significant association was found for presence of comorbidity (AOR = 1.08, 95% CI: 0.77-1.53) or female sex (AOR = 0.98, 95% CI: 0.69-1.37) in the adjusted analysis. The odds of mortality among COVID-19 cases not treated with oxygen therapy was 2.07 times the odds among cases receiving oxygen therapy (95% CI: 1.56-2.75, p<0.001), after adjusting for age, sex, and presence of underlying comorbid conditions. convalescent plasma was found to be significantly (OR: 0.5, 95% CI: 0.24-0.93, p=0.038) associated with reduction in mortality after adjustment for age, sex, and comorbidities.

**Table 1.**
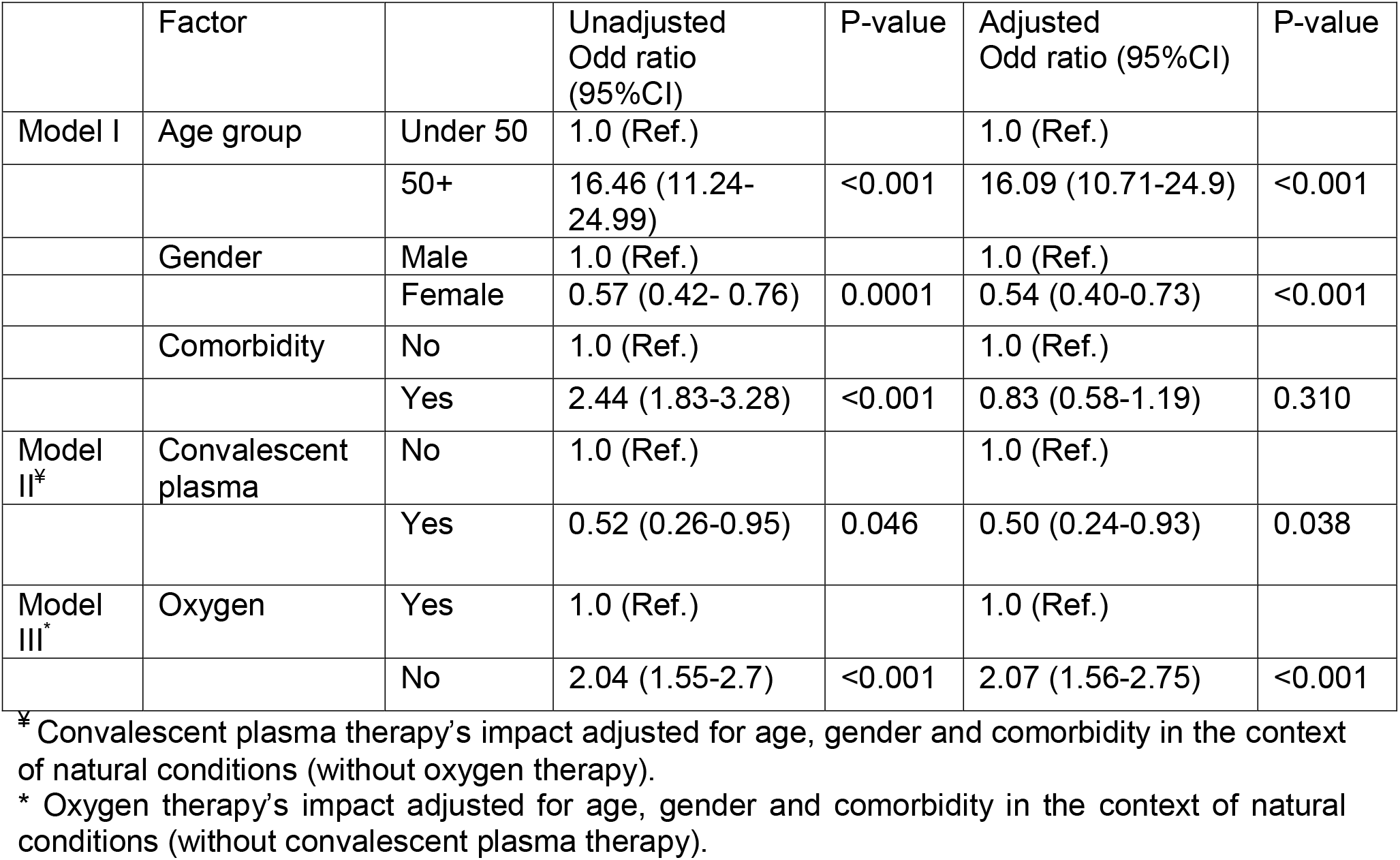
Results of logistic regression analysis to assess potential for reduced odds of mortality among cases receiving clinical intervention with oxygen therapy or convalescent plasma

## Discussion

Here we present data from a retrospective analysis of clinical and demographic characteristics of deceased COVID-19 cases in SSA. Deaths in the region were more frequently among individuals who self-reported or were tested postmortem compared to among cases identified through active monitoring. This ratio is notably different than that previously identified^7^ for the ratio of overall case detection, which was heavily skewed towards detection via active monitoring among all reported cases.

Deceased cases in Burkina Faso tended to be older with a higher proportion male, relative to all COVID-19 cases hospitalized for treatment. The data further suggests late or no care-seeking among the most severe cases that result in death. This is consistent with observations in other SSA countries. In Liberia, for instance, 11 of 18 reported deaths as of May 10 occurred in communities and only one occurred in a designated COVID-19 facility.^26^ Nigeria has also reported clusters of deaths among individuals who were not suspected of the disease^27,28^ suggesting that health seeking and access to care are relevant for reducing COVID-19 mortality.

Importantly, the data from Burkina Faso indicate that early detection, oxygen therapy as well as effective treatments are important to mitigating disease progression and preventing mortality. Using a synthetic case study population matched to data from West Africa, we demonstrate the potential effects of appropriate oxygen therapy and convalescent plasma in reducing odds of death among COVID-19 cases. Preliminary evidence has suggested that blood-related products, such as convalescent plasma, may attenuate COVID-19 disease severity or duration.^29^ Convalescent plasma was widely used in previous viral outbreaks ^25,30,31^ and the mechanism underlying its effectiveness is well described.^32,33^ For instance, during the 1918 pandemic of Spanish Flu, patients who received convalescent plasma therapy saw a reduced mortality rate, despite treatments being given to more severely ill patients.^25^ Similar results have been estimated for H1N1 flu treatment in smaller studies.^34^ Furthermore, the use of convalescent plasma as treatment in LMIC settings has precedent from the Ebola outbreak in West Africa, where mobile laboratories were used for the apheresis process.^31^

Of note, our investigation on convalescent plasma was implemented to reflect the range of uncertainty of its effect size (mortality risk difference of 10.5%, 95% CI: −4% to 25%) as observed in studies addressing convalescent plasma’s effectiveness against the Spanish flu. Further, well-designed studies will allow for better quantification of convalescent plasma effectiveness (i.e., with less uncertainty) and for informing treatment policies by providing clearer evidence of any risks versus benefits.

The results presented here are based on parameters informed by the use of oxygen and convalescent plasma on other respiratory diseases (2018 Spanish Flu) and hypoxia (respiratory and brain injury) disease, respectively, such that our findings should be interpreted with caution and revisited as empirical data from the clinical treatments provided to COVID-19 patients becomes increasingly available. Despite the limitations, highlighting and evaluating low-cost, easily scalable approaches for clinical management is critical to lower the CFR and to encourage timely care-seeking. This is particularly important in settings with significant undetected community transmission, so that individuals who develop COVID-like symptoms seek care early and are not deterred by the notion that the disease has no cure.

When considering clinical interventions, it is important to recognize that availability alone is not adequate to ensure proper clinical care. Reports on oxygen availability in LMICs have indicated that use is common but heterogeneous,^35^ with greater availability in more urban environments and at higher levels of care due to reduced access to pulse oximetry in more rural settings and at lower levels of care.^36-38^ In addition to requiring the necessary equipment, patients and caregivers must be willing to accept and administer the care respectively, which has been observed as a barrier^39^ such as due to perceived cost and lack of familiarity with the equipment which will need to be addressed during the COVID care response. This is concerning because if patients are escalated to higher levels, this introduces delays in care, and means that they will bring COVID with them to the hospital populations. In parallel to building treatment capacity, risk communication and community engagement^40^ will be essential to increasing awareness of its availability and affordability, so that people will actually seek care promptly. Critical cases who delay care-seeking would likely arrive too late for oxygen or other interventions to have much effect.

If oxygen therapy can reduce the rate of severe disease, and convalescent plasma can reduce progression and mortality, it may be possible to substantially reduce the impact of COVID^41^ without the need for critical care and respirators, which are more difficult to rapidly scale up capacity for than these treatments would likely be. Expanding access to oxygen treatment not only benefits COVID patients, but would likely also have long-term impact on child mortality due to pneumonia or all causes^42^; literature has shown risk reductions of about 20-60% in small-scale studies in LMIC.^42^ Investment in capacity and protocols for use could have significant and long-term implications on overall disease burden.

We provide the first study of data from deceased COVID-19 cases in the sub-Saharan Africa region. While absolute counts in SSA are considerably lower than in other regions globally, low-cost and readily scalable case management approaches remain important to reducing mortality due to COVID-19 in resource-constrained settings. Investing in capacity for oxygen therapy administration and apheresis, along with research infrastructure to assess effectiveness, could not only improve outcomes during the pandemic response but also have long-term implications for health systems.

## Data Availability

Data are presented in tables and figures within the text.

## Authors’ contributions

LAS, BH, BA, EAW and ALO designed the study. LAS, NN, MVG, KD, MK, AG, IV, HT, BWB, and HH collected data for the linelist or at the Center Hospitalier Universitaire de Tengandogo in Ouagadougou. ALO conducted the data simulation. LAS and ALO conducted the data analysis. LAS and ALO wrote the first draft of the manuscript. All authors contributed intellectually and made contributions to the manuscript text.

## Competing interests

The authors declare no competing interests.

## Notes

### Competing Interest Statement

The authors have declared no competing interest.

